# The association between HDL-C and stroke in people over 40 years of age: A NHANES cross-sectional study

**DOI:** 10.1101/2024.04.07.24305464

**Authors:** Kevin Patel

**Affiliations:** Independent researcher, UNITED STATES

## Abstract

1.

**Background:** Previously, studies have indicated that high-density cholesterol (HDL-C) levels are associated with lower risk of cardiovascular disease. However, this has been challenged in recent years.

**Method:** This cross-sectional study was conducted on data from the National Health and Nutrition Examination Survey 2011-2020. There were total of 15232 out of 45462 individuals who were age >40 years old, not missing HDL-C levels and not missing stroke data. The independent variable was HDL-Cholesterol. The outcome variable was stroke. We used multivariate logistic regression models to assess the relationship between stroke and HDL-C in three models consisting of the following explanatory variables: 1. HDL-C alone (unadjusted model), 2. HDL-C and non-modifiable risk factors (age, sex and race), 3. HDL-C, non-modifiable, and modifiable risk factors [hypertension, diabetes and smoking status] risk factors. We also conducted analyses stratified by sex and race.

**Results:** HDL-C categories 40-59 mg/dl (unadjusted model- 0.759, CI: 0.639, 0.901; non-modifiable risk factors adjusted- 0.668, CI: 0.558, 0.798) and 60-79 mg/dl (unadjusted model- 0.733, CI: 0.598, 0.898; non-modifiable risk factors adjusted- 0.565, CI: 0.455, 0.701) had significantly lower risk of stroke relative to HDL-C category <40 mg/dl. But when adjusted for modifiable risk factors, risk of stroke HDL-C categories 40-59 mg/dl and 60-79 mg/dl categories were not significantly different from the HDL-C <40 mg/dl category. The risk of stroke HDL-C categories 80-89 mg/dl and >90 mg/dl categories were not significantly different from the HDL-C <40 mg/dl category in all models. In stratified analyses, HDL-C was significantly inversely associated with the risk of stroke in male gender and in non-Hispanic White race.

**Conclusion:** Our study reveals various inverse relationship between HDL-C and risk of stroke. These findings suggest that relevant levels of HDL-C may be beneficial in preventing stroke in the absence of modifiable risk-factors.

## 2. Introduction

Stroke is one of the leading cause of death and disability in United States. Each year about 795,000 people have strokes.^1^ About 87% of all strokes are ischemic strokes, and 13% are hemorrhagic strokes.^1^ In 2021, 162,890 people died due to stroke in US.^2^ Certain condition such as high blood pressure, diabetes mellitus, heart disease (like atrial fibrillation), previous stroke, dyslipidemia, obesity and smoking.^1^ However, targeted intervention to reduce BP, blood sugar and blood cholesterol levels can substantially reduce the burden of stroke.^3^ High-Density Lipoprotein (HDL) is consider a good type of lipoprotein. It carries cholesterol from the peripheral blood to the liver to be excreted out of the body. In addition, HDL-C exhibit beneficial effect on platelet function, endothelial function, coagulation, and inflammation. Also, it has antioxidant effect, all of which play key role in preventing atherosclerosis.^4^ Despite these benefits, studies researching association between HDL-C and stroke remains limited and controversial.

Many previous studies showed that higher HDL-C was associated with lower risk of stroke.^5^ Some studies found that there is no association between HDL-C and cholesterol.^6,7^ EUROSTROKE study showed that in men HDL-C levels slowly trends towards lower stroke risk, but it increases risk of non-fatal stroke and hemorrhagic stroke in women.^8^ Wang et al also showed that higher LDL-C lowers risk of hemorrhagic stroke and higher HDL-C increases risk of hemorrhagic stroke.^9^ A recent meta-analyses reported that higher levels of HDL-C may be associated with increased risk of stroke.^10^ Due to these various associations, the treatment of increasing HDL-C is being challenged.

Very few studies have reported the relationship between high density lipoprotein and risk of stroke with different sex and races. We aim to examine the relationship between HDL-C and risk of stroke in people older than 40 years of age using the National Health and Nutritional Examination Survey (NHANES 2011-2020). Our findings can aide development of preventive strategies of stroke based on sex and race.

## 3. Methods

### a. Study design, population, and data collection

The NHANES is a population-based national survey of the United States that collects health and nutrition data every two years. After standardized home interview, physical examination and biological samples are collected in mobile examination centers. The NHANES is a cross sectional study design that aims to produce nationally representative sample of US population. We obtained HDL-C and stroke data from NHANES 2011 and 2020 (henceforth reffered to as the 2011-2020 cycle). Overall, 45,462 people participated in NHANES 2011-2020. There was total OF 17,169 people over the age of 40 years. We excluded people with missing stroke data (n=27) and people with missing HDL-C (n=1937). Our final analysis included 15,232 participants.

### b. Definition of stroke

In NHANES, participants were asked, “Has a doctor or other health professional ever told you that you had stroke?” In the US, self-reported stroke is generally accurate among the general population, and it has been used in many previous studies. Stroke is used as dependent variable.

### c. Definition of HDL-C

HDL-C was measured using direct immunoassay method. There were no changes to the lab method or lab site for the NHANES 2011-2020 cycle. In the 2011-2012 cycle, the three analytes were measured on the Roche modular P chemistry analyzer. In the 2013-2020 cycle, the three analytes were measured on the Roche modular P and Roche Cobas 6000 chemistry analyzers.

We used HDL-C as independent variable. First, it was used as a continuous variable to assess risk of stroke. We also analyzed relationship between them, when HDL-C was used as a categorical variable. We divided the HDL-C levels into five categories: HDL-C <40 mg/dl, 40– 59 mg/dl, 60–79 mg/dl, 80–89 mg/dl, and ≥90 mg/dl.

### d. Covariates

As covariates, we included age, body mass index (BMI), large density lipoprotein cholesterol (LDL-C), sex, race, smoking status, diabetes and hypertension. Race was categorized into Mexican American, other Hispanic, non-Hispanic White, non-Hispanic Black, and other races. Smoking status was categorized in current smokers (people who are currently smoking), and non-smokers (people who never smoked or previous smokers). BMI and LDL-C data were collected by mobile examination centers. Hypertension and diabetes were self-reported during the questionnaire.

### e. Statistical analysis

All statistical analysis were conducted using SAS studio software. A p-value of <0.05 was considered statistically significant. Continuous variables were presented as means ± standard deviation (SD), and categorical variables were presented as total and percentage. Continuous variables were checked for normality first, then one way analysis of variance test was done to check statistical significance against categorical variables. Chi-square test was used to check significance between categorical variables. Generalized logistic regression was used to examine the association between stroke and HDL-C. We used 3 models: 1. Unadjusted model that solely included HDL-C as independent variable, 2. HDL-C and adjusting for non-modifiable risk factors (age, sex and race) were adjusted, 3. HDL-C and adjusting for non-modifiable risk factors and modifiable risk factors (smoking status, diabetes, hypertension, BMI and LDL-C) were adjusted. We first used HDL-C as a continuous variable in our analyses. To further explore the relationship between stroke and HDL-C, we also conducted analyses with HDL-C levels categorized into five groups: HDL-C <40 mg/dl, 40–59 mg/dl, 60–79 mg/dl, 80–89 mg/dl, and ≥90 mg/dl. Subgroup analysis stratified by race and sex were also performed.

## 4. Results

There were total of 15,232 eligible participants for our study. Table 1 shows the sample baseline characteristics. There was significant difference in the baseline categories of all covariates among the HDL-C groups. Male participants were more in lower HDL categories, and female participants tend to be more in higher HDL-C categories. The higher the HDL-C categories, the lower the BMI. Lowest HDL-C groups are more likely to develop diabetes and hypertension.

**Table 1.**
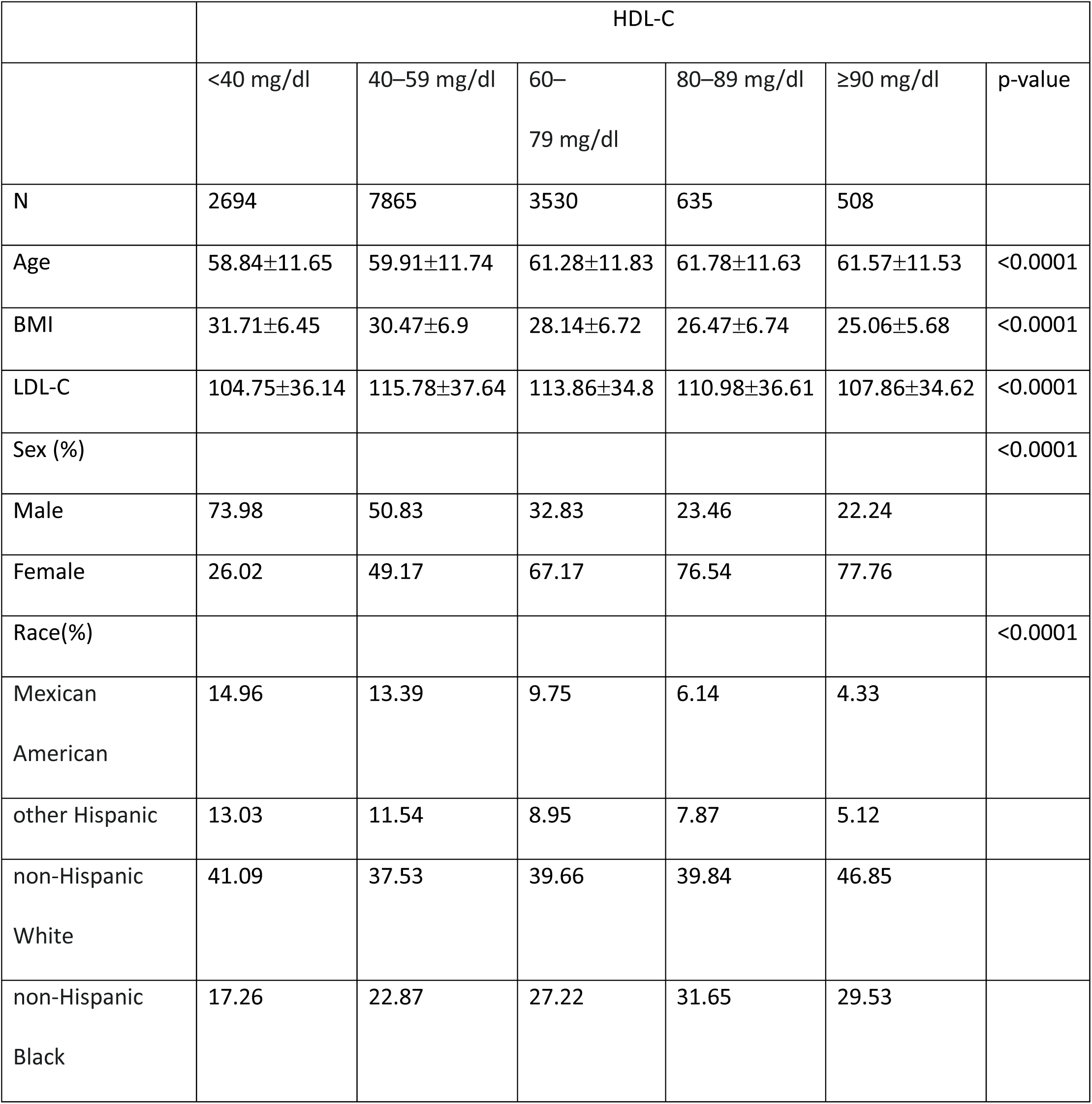
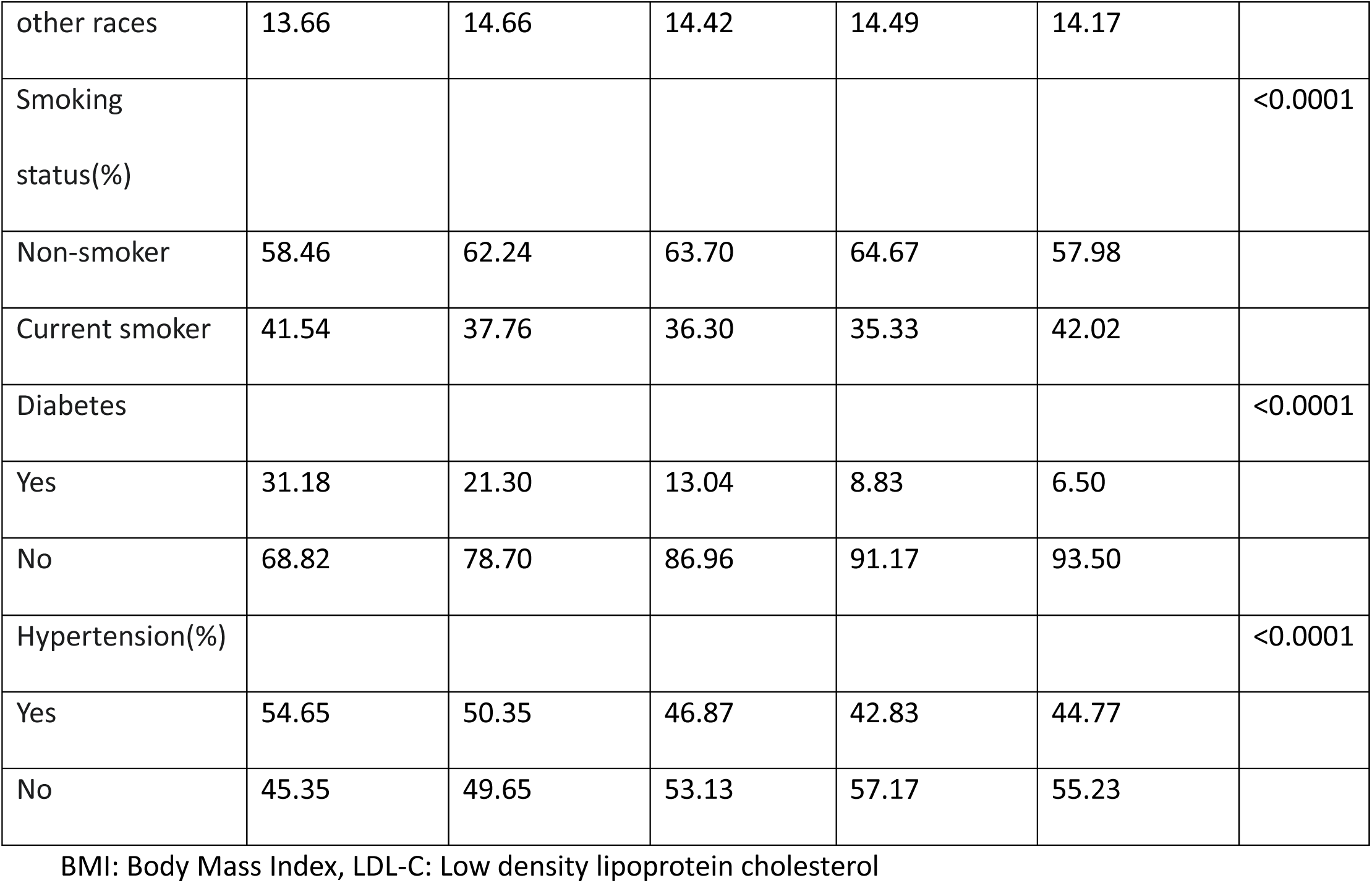
Baseline characteristics of participants.

Table 2 shows the results of generalized multiple regression analysis. In the unadjusted model (Odds Ratio (OR) for continuous HDL-C= 0.996, 95% CI: 0.991- 1.0, p-value- 0.0361) and in the model adjusted for non-modifiable risk factors (OR= 0.99, 95% CI: 0.985- 0.994, p-value <0.0001), HDL-C was inversely associated with the risk of stroke. However, in the model adjusted for non-modifiable and modifiable risk factors (OR= 0.994, 95% CI: 0.0986- 1.003, p-value- 0.1711), HDL-C was not associated with the risk of stroke. In the models for HDL-C as a categorical variable (reference category = <40 mg/dl), HDL-C levels between 40- 59 mg/dl was inversely associated with risk of stroke in unadjusted model (OR= 0.759, 95% CI: 0.639- 0.901, p-value- 0.0017) and in the model adjusted for non-modifiable risk factors (OR= 0.668, 95% CI: 0.558-0.798, p-value <0.0001). In HDL-C category of 60-79 mg/dl, the risk of stroke is reduced by 27% in unadjusted model (OR= 0.733, 95% CI: 0.598- 0.898, p-value 0.0027) and by 44% in model adjusted for non-modifiable risk factors (OR= 0.565, CI: 0.455- 0.701, p-value <0.0001). In the model adjusted for non-modifiable and modifiable risk factors, HDL-C categories of 40-59 mg/dl (OR= 0.735, 95% CI: 0.517- 1.045, p-value 0.866) and 60-79 mg/dl (OR= 0.713, 95% CI: 0.466- 1.090, p-value 0.1186) were not significantly associated with risk of stroke. However, in models adjusted for non-modifiable risk factors, the highest HDL-C category was significantly associated with risk of stroke.

**Table 2.**
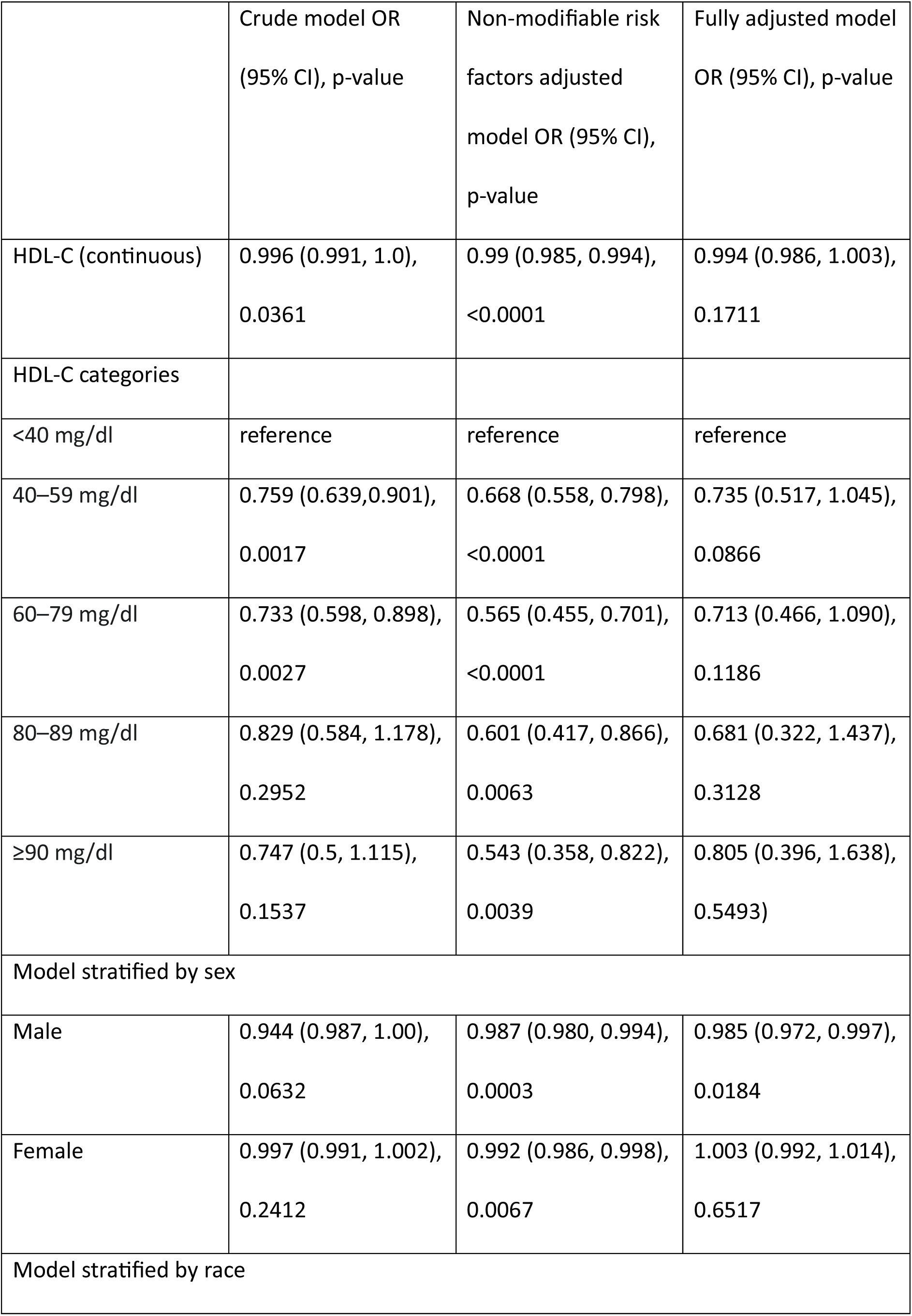
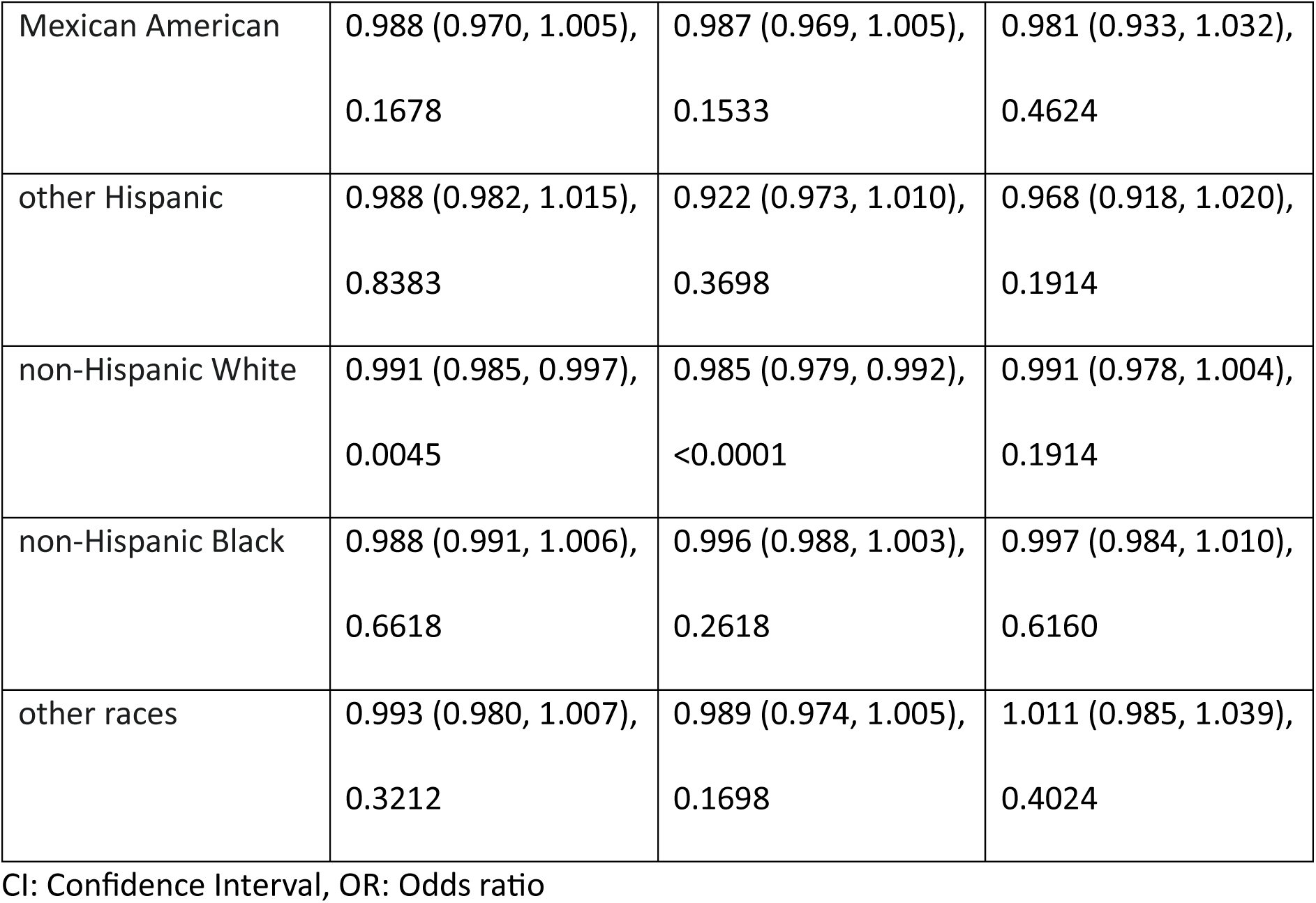
The association between High density lipoprotein cholesterol (HDL-C) and stroke.

Stratified by sex, HDL-C was not significantly associated with risk of stroke in unadjusted models for both males (OR= 0.944, 95% CI: 0.987- 1.0, p-value 0.0632) and females (OR=0.997, 95% CI: 0.991- 1.002, p-value – 0.2412). However, in models adjusted for non-modifiable risk factors and fully adjusted models in men, HDL-C was significantly associated with lower risk of stroke. In contrast, in females, HDL-C was significantly associated with stroke only in models adjusted for non-modifiable risk factors. When stratified by race, HDL-C was significantly associated with lower risk of stroke in other Hispanic white race in unadjusted model (OR= 0.991, 95% CI: 0.985- 0.997, p-value 0.0045) and in the model adjusted for non-modifiable risk factors (OR= 0.985, 95% CI: 0.979- 0.992, p-value <0.0001). However, in the fully adjusted model for non-Hispanic white race, HDL-C was not significantly associated with lower risk of stroke. All the other subgroups of race did not show any significance association between stroke and HDL-C.

## 5. Discussion

In our study, we found a several negative associations between risk of stroke and HDL-C. HDL-C groups of 40-59 mg/dl and 60-79 mg/dl were negatively associated with the risk of stroke. But interestingly, when we adjusted for comorbidities like diabetes, hypertension and smoking, there was no significant association between them. All the models including subgroup analysis, only male subgroup was negatively associated with risk of stroke and HDL-C, when we adjusted for comorbidities. In subgroup analysis, male and non-Hispanic white race were the only once negatively associated with risk of stroke in unadjusted and non-modifiable risk factors adjusted models. These findings suggest that HDL-C can be protect against stroke in the absence of comorbidities.

Even though relationship between HDL-C and stroke has been studied previously, but it has been always controversial. Multiple previous studies have reported lower risk of stroke with adequate level of HDL-C.^5,11,12^ Recent cohort study (Vitturi et al) supported the conclusion that HDL-C reduces the risk of stroke.^13^ EUROSTROKE study revealed no association between risk of stroke and HDL-C.^8^ Recent prospective study from 2022 revealed an U-shaped relationship between HDL-C and stroke.^14^ In our study, we found protective U-shaped relationship. In the absence of comorbidities, HDL-C levels of 40-79 mg/dl reduced the risk of stroke, while <40 mg/dl and >79 mg/dl did not. When we adjusted for modifiable risk factors of stroke i.e., diabetes, hypertension, LDL-C (hyperlipidemia) and BMI (obesity), we found no associated between HDL-C and stroke. Previous studies have shown that these risk factors increase the risk of stroke substantially.^15^ One of the reasons for no association between HDL-C and stroke in the presence of modifiable risk factors can be the size of HDL-C particle. Prospective nested case control study from Japan considered HDL-C particle size into account for risk of stroke, and they found that large HDL-C particles were not associated with risk of stroke compared to small and medium sized HDL-C particle.^14^ Previous study found that HDL particle size may be a better marker of residual risk rather than chemically measured HDL-C.^16^ Future studies need to consider how does the distribution of HDL-C particle size impacts the risk of stroke. Also, the effect of comorbidities like hypertension and diabetes on the HDL-C particle size.

In our study, we found that HDL-C and risk of stroke were significantly associated in men. However, in women, relationship was not significant between them. One of the reasons can be the lower estrogen levels in postmenopausal women, which in turn lowers HDL-C levels.^17^ Study from Women’s Health Initiative (WHI) in 2002 showed that hormone replacement drugs has more detrimental effect on women than beneficial, so recent use of these drugs has been reduced.^18^ These, in turn, reduced estrogen levels and HDL-C. There has not been much research on how ethnicity effects the association between HDL-C and stroke. Our study revealed that only non-Hispanic white race showed a significant relationship in the absence of comorbidities. These factors can be explained by socioeconomic factors and genetic factors. Further prospective cohort studies need to done to evaluate the role of HDL-C in stroke prevention.

This study can be subject to some limitations. First, NHANES uses widely standardized tools to measure the laboratory values, but it can be subject to measurement bias. This can lead to misclassification bias in the overall sample. However, they have taken steps to prevent such biases and usually their clinical data is considered quite accurate and reliable. Second, NHANES is a cross-sectional survey, so it presents its own challenges as it does not allow us for causal assessment. Third, NHANES does not distinguish between hemorrhagic and ischemic stroke. In future, studies should take into the consider the type of stroke. Fourth, NHANES and generally do present some challenge recently, we do not have standardized way to measure the HDL-C particle size, which plays important role in pathogenesis of stroke. On the other hand, our study has some strengths too. The large NHANES study population is representative of the entire US population, and interviews and examinations are carried out by the trained personnel. The large sample size also allows us to do stratified analyses and adjustment for multiple confounders and effect modifiers. Also, we were able to assess the relationship between risk of stroke and HDL-C based the type of risk factors i.e., non-modifiable (age, sex and race) and modifiable (smoking status, hypertension, diabetes, BMI and LDL-C).

## Summary

We found several negative associations between HDL-C and stroke. HDL-C is negatively associated with risk of stroke in unadjusted and non-modifiable risk factors (age, sex and race) adjusted models. However, when we adjusted for modifiable risk factors (hypertension, diabetes, obesity and LDL-C), there was no significant association. This was true for both HDL-C as continuous and categorical variable. When HDL-C was used as categorical variable, only 40-59 mg/dl and 60-79 mg/dl categories were associated with reduced risk of stroke in unadjusted and non-modifiable risk factors adjusted models. 80-89 mg/dl and >90 mg/dl categories were not significant in any models. When we stratified by sex, only men had reduced risk of stroke. Even when we adjusted for modifiable risk factors, men had reduced risk of stroke. In race stratification, only non-Hispanic white race was significantly associated with risk of stroke in unadjusted and non-modifiable risk factors adjusted models.

## Data Availability

All datasets are available on the National Health and Nutrition Examination Survey website.

